# Cardiorespiratory demands of firearms training instruction and 15m shuttle tests in British law enforcement

**DOI:** 10.1101/2024.02.27.24303347

**Authors:** Joseph Warwick, Sophie Cooper, Flaminia Ronca

## Abstract

**Objectives:** Law enforcement agencies require minimum fitness standards to safeguard their officers and training staff. Firearms instructors (FI) are expected to maintain the same standards as their operational counterparts. This study aimed to quantify the daily physiological demands placed on FI.

**Methods:** 19 FI (45 ± 5 years) completed occupational tasks whilst wearing heart rate (HR) monitors for a minimum 10 days. Maximal oxygen consumption (VO_2_max) testing was conducted on FI during a treadmill test (TT) and a multistage shuttle test (ST). Linear regression models were used to model the relationship between VO_2_ and HR throughout the TT. This model was applied to HR data from occupational tasks to infer oxygen consumption. Repeated Measures ANOVAs were used to compare time spent in VO_2_max equivalent zones throughout.

**Results:** The VO_2_max achieved during ST (45.1 ± 5.6 ml/kg/min) was significantly higher than TT (39 ± 3 ml/kg/min) (p = 0.014). Time to exhaustion (TTE) was sooner on ST (06:26 min) compared to TT (13:16 min) (p < .001). FI spent ∼85% of occupational time with an oxygen demand ≤20 ml/kg/min (p < .005). The most intense occupational tasks saw FI achieve VO_2_max ≥30 ml/kg/min, but <40 ml/kg/min.

**Conclusion:** Using ST to assess cardiorespiratory fitness resulted in a quicker TTE and a higher VO_2_max. Predominantly, FI occupational tasks are low intensity with sporadic exposures requiring a VO_2_max of >40 ml/kg/min. To safeguard FI from occupational-related cardiorespiratory or long-term health issues, it is intuitive to suggest fitness standards should exceed a VO_2_max of 40 ml/kg/min.

## INTRODUCTION

In recent years, a significant portion of public service personnel, including law enforcement officers [1],firefighters [2,3], paramedics [4], and military personnel [5], have globally shown a reduced level of cardiorespiratory fitness, increased body mass index (BMI) and increased fat free mass[1–4]. This is partly due to changes in recruitment standards and the concurrent implementation of human rights legislation with regard to physical standards for employment in 2010 [1,6] The legislation within the United Kingdom states that employers must be able to defend physical standards as ‘reasonably necessary to the safe and efficient performance of the job’ and ‘to employ the standards, tests and cut-off scores that adequately capture the minimum requirements to do a job safely and efficiently’ [6].

Throughout their careers, specialist law enforcement officers who train for firearms operations develop high levels of skill, making employee retention an important focus for this population. The more experienced and skilled firearms officers become a strong asset to their forces, contributing to the development and education of new recruits as firearms instructors (FI), sometimes even after retirement from operational duties. Key elements of an instructor’s responsibilities include live range teaching and active role play, some of which can require significant physical effort, all while also instructing and teaching. Supporting the physical fitness of this group is therefore highly relevant to their occupational performance, their ability to instruct while under exertion, and to enable their retention. Heart rate and stress-related heart rate variability (HRV); particularly, HRV-derived sympathetic responses have been shown to directly impact shooting accuracy and success [7–9], with the level of expertise directly impacting cognitive control and timing of the shot with the cardiac rhythm [10,11]. A reduced heart rate during the initiation of shooting also determines the shooter’s ability to control shot timing and accuracy [11,12].

Cardiovascular fitness is therefore essential to enable FI to maintain a good level of performance, and to withstand the occupational demands of role play and instruction during officer training, while also safeguarding their own health.

The maximal oxygen carrying capacity (VO_2_max), is the gold standard measure of cardiovascular and pulmonary fitness and reflects one’s ability to sustain specific levels of physical exertion[13,14]. Cardiorespiratory fitness testing has historically played an integral part in officer and FI recruitment and is still a key pre-requisite for enrolment and annual revalidation in many law enforcement settings. The minimum requirements of fitness should be determined based on the demands of operational tasks, to ensure that officers and FI are able to fulfil their duties without incurring unnecessary fatigue. Therefore, the aim of this study was to assess the cardiorespiratory demands placed on FI during law enforcement officer firearms training, including exertion undertaken during instruction and job-related fitness testing.

## METHODS

### Study Design

This study was designed as an observational cohort study with Firearms Instructors (FI)voluntarily recruited through internal emails from a British law enforcement service. This was sent via line managers in their workforce during January and February 2021. A total of 19 FI aged 45 ± 5 years volunteered to take part (18 male, 1 female). One participant was non-operational and 18 were operational law enforcement officers, of these, 16 (1 female) were armed vehicle response officers (AVRO) and 2 were firearms officers (FO). All participants signed a written informed consent form and a physical activity readiness questionnaire (PARQ) before participating. The study was approved by UCL’s Ethics Committee (Project ID number: 13985/004) in line with the declaration of Helsinki.

Anthropometric measurements and body composition were taken using a multi-frequency (1kHz/5kHz/50kHz/250kHz/500kHz/1000kHz) electrical bioimpedance with a calibrated 8-segment analyser, the Tanita MC980 PLUS MA (Tanita Cooperation, Tokyo, Japan) which has an accuracy of 0.1kg/0.1% and in compliance with MDD 93/42/ECC directive, certified for medical use within Europe [15,16]. Participants were advised to refrain from caffeine consumption and exercise for 24 hours before arriving at the laboratory and also arrive hydrated. In a randomised order, participants then completed two VO_2_max tests. One VO_2_max test was conducted on a treadmill (TT) using the incremental Bruce treadmill protocol [17,18], commonly considered the gold standard VO_2_max test to evaluate maximal oxygen consumption. The second VO_2_max test was a 15-meter shuttle test (ST) [19], which consisted of the volunteers’ annual fitness revalidation test and is often used as a field-based indirect measure of VO_2_max in British law enforcement settings. After this, all participants recorded their heart rate (HR) for a minimum of 10 days during occupational firearms instruction activities. This included but was not limited to classroom theoretical teaching, live firearms instruction on the firing range, external open countryside assailant search and restraint, assailant restraint and arrest instruction, and vehicle stop and restraint instruction. The HR data for this period was collected via a polar chest strap HR monitor (H9, Polar, Kempele, Finland) through the Polar Beat app.

### Treadmill VO_2_max procedure

The lab-based VO_2_max test was conducted on a treadmill (h/p/cosmos, Nussdorf, Germany) using the incremental Bruce treadmill protocol and breath-by-breath analysis with a Vyntus CPX Metabolic Cart (Vyaire Medical, Chicago, USA) [17,18,20–22]. The incremental protocol commences with a three-minute warm-up walking at 2.7 km/h with no incline. Every three minutes thereafter, the speed and incline of the treadmill are increased, and participants are verbally encouraged to push themselves to maximal exertion. The test is terminated at volitional exhaustion, at which point participants walk on no incline at 2.7 km/h until their HR recovered to under 120 bpm. A test was considered maximal if the participant reached their predicted maximal heart rate (220-age), if RER > 1.15 and if a plateau was observed in the VO_2_ data. All participants in this study reached these criteria. The anaerobic threshold was determined using the v-slope method.

### 15m Shuttle test VO_2_max procedure

In a randomised order, participants also completed a 15m multistage shuttle test (ST) while wearing a portable breath-by-breath analyser (Metamax 3B, Cortex, Leipzig, Germany) [22–24]. This was completed indoors to alleviate external environmental influences and required individuals to repeatedly run back and forth between two markers 15 meters apart in time with an electronic beep. The ST requires participants to run progressively faster, level one allowing 6.8 seconds per shuttle, with incremental increases in pace to >4 seconds per shuttle in the later stages [23,25]. All participants completed to volitional exhaustion with verbal encouragement.

### Data Processing Treadmill Test

Breath-by-breath VO_2_ (ml/kg/min) and HR (bpm) were smoothed to every 7 breaths, standardising the data across both testing protocols, using the Vyaire interface and then extracted from the software. To estimate oxygen requirements during occupation-related tasks, a correlation factor was calculated from the treadmill test. This was achieved by extracting the correlation equation from a linear regression plot comparing HR against VO_2_ for data collected during the treadmill test.

### Occupation-related task data

Occupation-related data was collected using the Polar coach module, the most strenuous session per participant was then downloaded and analysed using the Pandas module for Python (Python Software Foundation. Python Language Reference, version 3.7. Available at http://www.python.org). HR data from the entirety of the most strenuous session was correlated to a VO_2_ equivalent to provide an estimate of oxygen requirements throughout the task. Data was then analysed by categorising it into two samples: the highest recorded value for HR and VO_2_ in each minute of the tests, and the highest recorded value for VO_2_ in each HR category. Categories were recorded in 10bpm increments from 50 to 190.

### Shuttle test data

Breath-by-breath VO_2_ (ml/kg/min) and HR (bpm) were extracted from the software and then also smoothed to every 7 breaths using Python. Shuttle timings were used to label the data instance to the appropriate ‘Stage: Shuttle’ category. Maximum values for HR and VO_2_ for each category, for each participant, were then calculated using pre-built tools from the Pandas (version 1.04) module for Python.

### Statistical analysis

The data was analysed using the statistical software SPSS 26 (IBM, New York, USA), with alpha set at <0.05. All data were normally distributed. Linear regression models were used to model the relationship between VO_2_ and HR for each participant throughout the treadmill VO_2_max test, as described above. The model was then applied to the polar HR data collected during job-related tasks to infer oxygen consumption during working hours. Paired t-tests were used to compare VO_2_max and maximum HR between the treadmill test and shuttle test. Repeated Measures ANOVAs with Bonferroni post-hoc were used to compare time spent in VO_2_max equivalent zones within participants. A Mauchley’s test of sphericity was initially carried out to confirm the assumption required to run an ANOVA.

## RESULTS

### Participant Demographics

Participant fitness and anthropometrics are shown in Table 1.The mean BMI was 28.1 ± 2.9, with 3 participants in the normal range of 18.5 to 24.9, 11 participants classified overweight between 25 and 29.9 and 5 participants classified obese at >30 but <35. No participants were classified as morbidly obese wih a BMI >35. Regarding body fat percentage those under 40 years old (n = 5) had a range between 17% and 26%, those 40 to 49 (n = 11) range from 13.5% to 27.7% and those 50+ (n = 4) range from 19% to 29.1%. Since the female participant’s data did not differ from the mean values of the cohort, male and female data was combined into one group to safeguard the anonymity of participants’ outcomes.

**Table 1.**
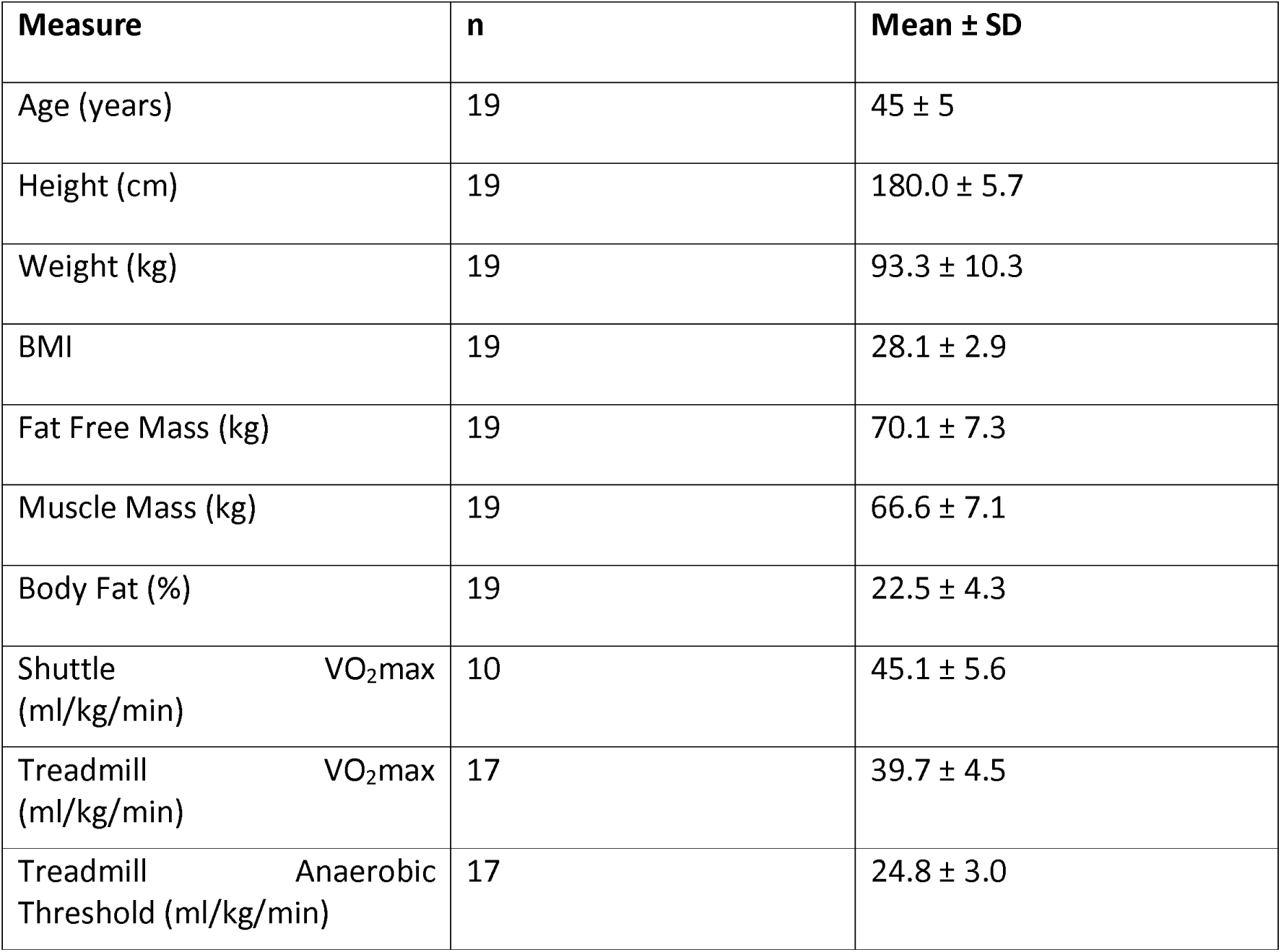
Mean ± SD values for anthropometric and fitness data.

### Comparison between the treadmill test and the 15m shuttle test

Ten participants completed both the TT and the ST, data from two ST was excluded, one due to hardware failure towards the final stages of the ST and the other due to software failure after the ST. The data presented in this section only relates to those eight participants who provided completed data on both tests (male = 7; age 43 ± 7 years; height 177.4 ± 5.8 cm; weight 91.1 ± 9.4 kg). The VO_2_max achieved during the ST was significantly higher (p = 0.014) than that achieved during the TT (45.1 ± 5.6 ml/kg/min and 39 ± 3 ml/kg/min respectively) (Fig 1a). There was no correlation between VO_2_max measured during the ST and VO_2_max measured during the TT (Fig 1b).

**Fig 1.**
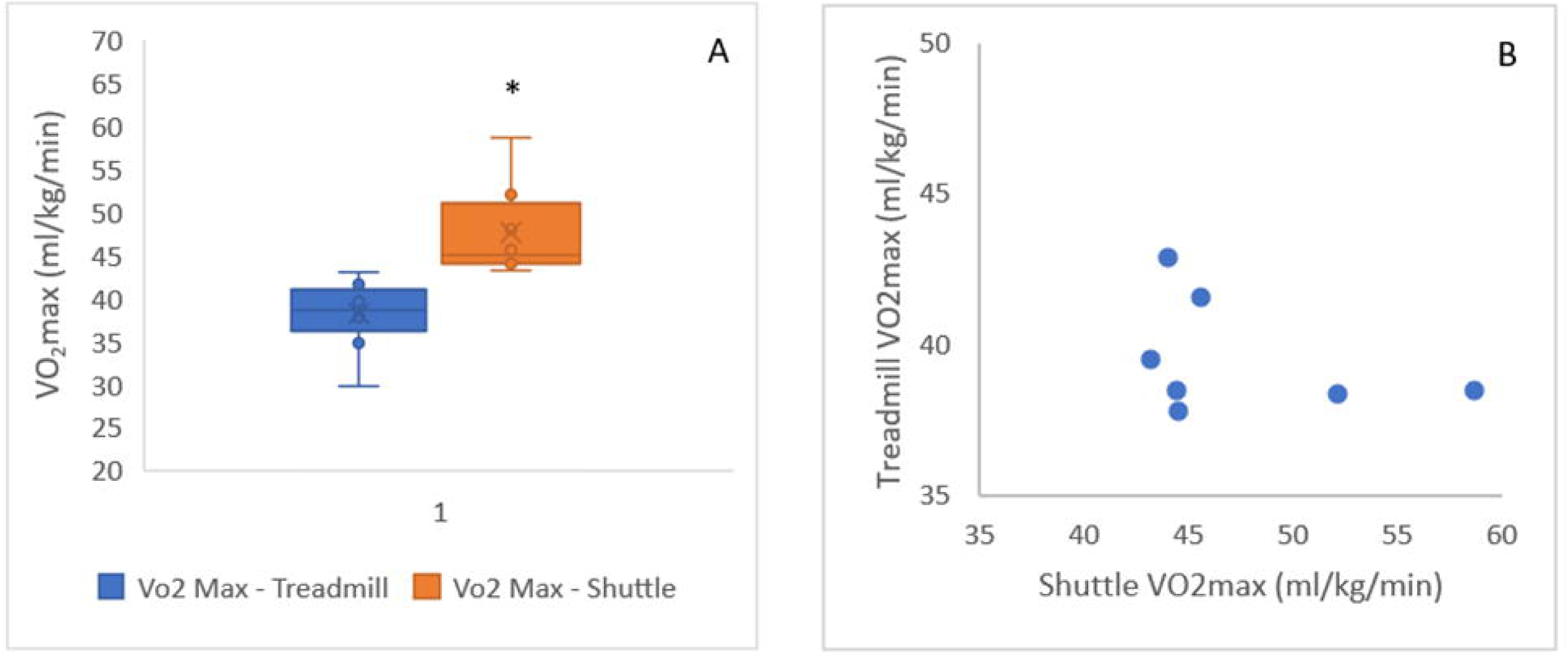
Comparison between the treadmill test and the 15m shuttle test. (A) Median ± IQR VO_2_max measured through the treadmill test and the shuttle test. * Significant difference between tests (p = .014). (B) Relationship between VO_2_max measured through either test.

All participants reached exhaustion sooner on the ST, with a total average duration of 13:16 min on the TT and 06:26 min on the ST (p < .001). Both heart rate and VO_2_ were higher throughout the ST compared to the TT (p < .001) from minute 2 to test termination (Figure 1). However, when comparing VO_2_ to HR zones (Fig 2) the difference between tests was not statistically significant(p = 0.068).

**Fig 2.**
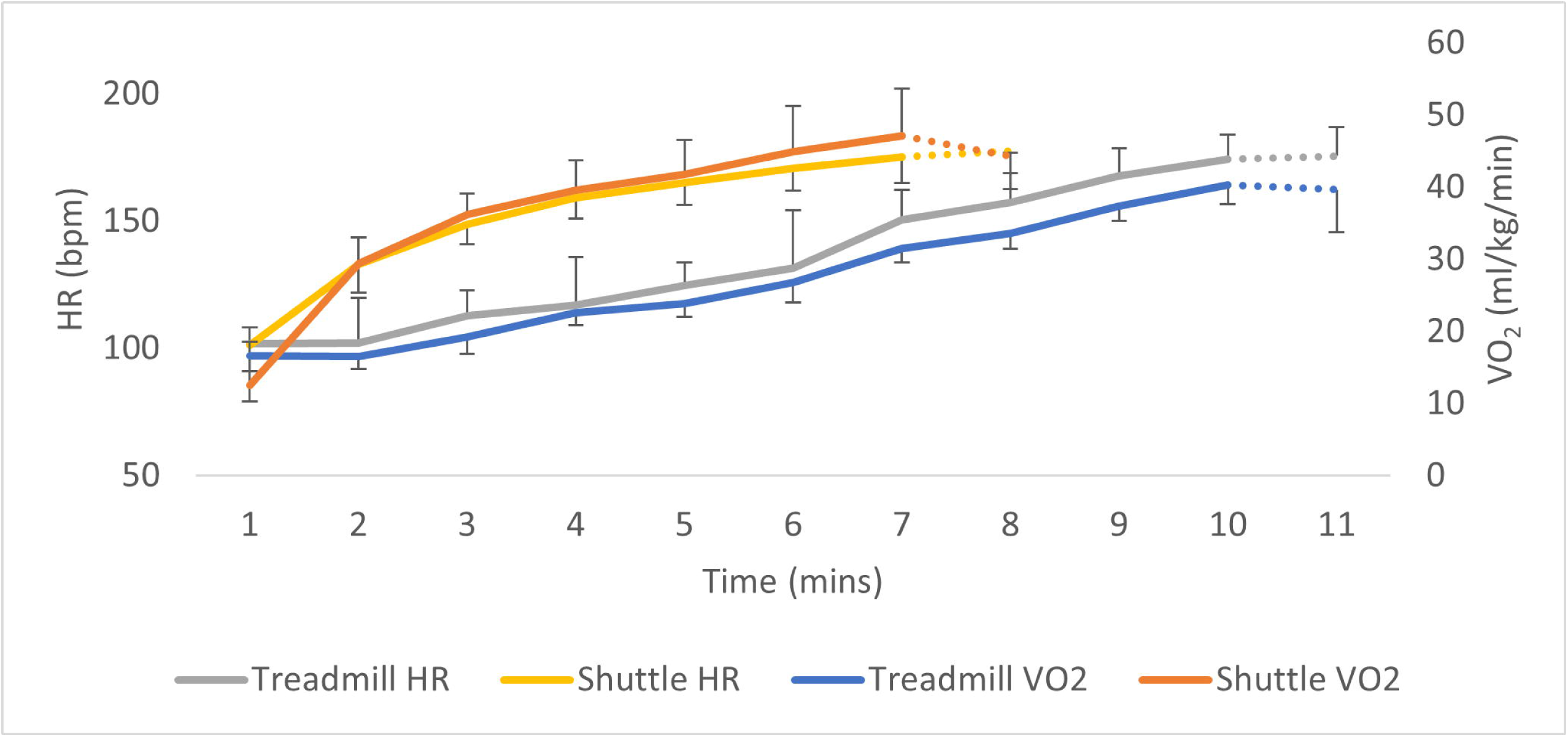
Mean ± SD heart rate and VO_2_ during the treadmill test compared to the shuttle test. (Note: the dotted lines denote incomplete data, where only participants who reached higher stages are included)

Despite a difference in the rate of increase of both VO_2_ and HR by time between tests (Figure 2), when comparing the rise in increase of VO_2_ concerning HR zones the difference was not statistically significant (p=0.068) (Fig 3).

**Fig 3.**
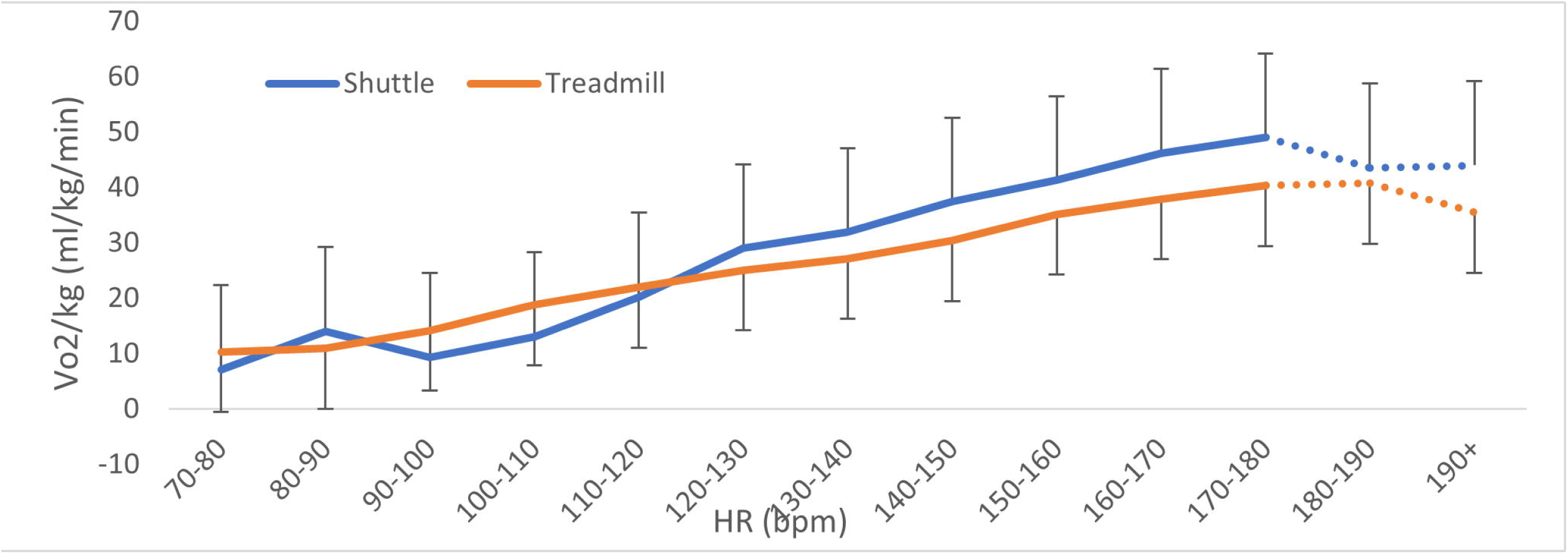
Mean ± SD rise in VO_2_ by heart rate zone during the shuttle test and treadmill test. (Note: the dotted lines denote incomplete data, where data for heart rate zones higher than 170-180 bpm include a subsample of participants whose hearts reached higher values)

The comparison of end-of-stage VO_2_max during the 15m shuttle test with previously studied data has shown observable differences Table 2.

**Table 2.**
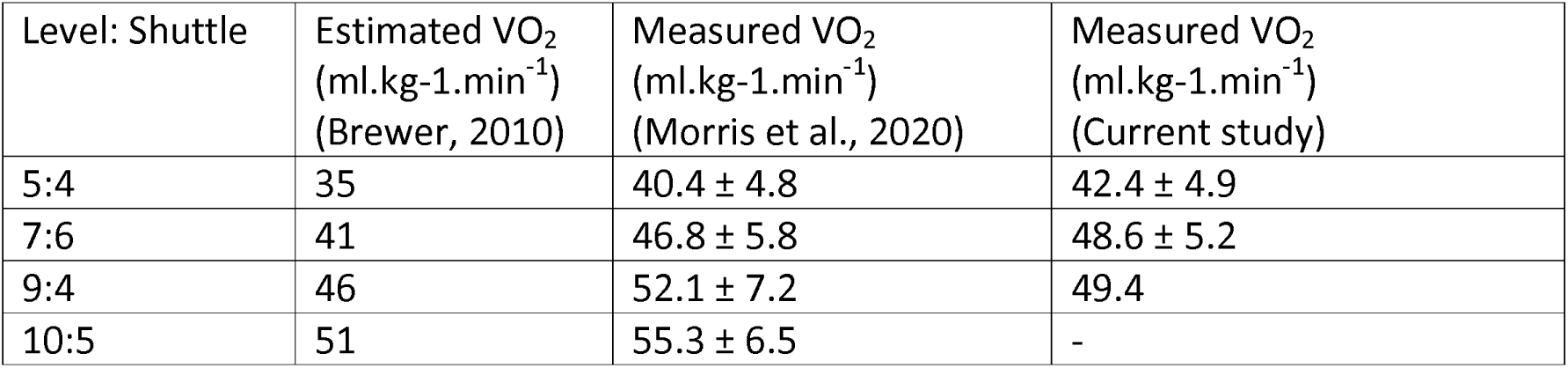
Estimated and measured VO_2_ equivalents per each end stage of the 15m shuttle test, based on findings from different papers.

| Level: Shuttle | Estimated VO<sub>2</sub><br>(ml.kg-1.min<sup>-1</sup>)<br>(Brewer, 2010) | Measured VO<sub>2</sub><br>(ml.kg-1.min<sup>-1</sup>)<br>(Morris et al., 2020) | Measured VO<sub>2</sub><br>(ml.kg-1.min<sup>-1</sup>)<br>(Current study) |
| --- | --- | --- | --- |
| 5:4 | 35 | 40.4 ± 4.8 | 42.4 ± 4.9 |
| 7:6 | 41 | 46.8 ± 5.8 | 48.6 ± 5.2 |
| 9:4 | 46 | 52.1 ± 7.2 | 49.4 |
| 10:5 | 51 | 55.3 ± 6.5 | - |

### Occupation-related physical exertion

Heart rate data was collected during occupational tasks for a total of 18 participants (17 male), with an average of 16 ± 9 instructional days recorded per individual, yielding an average total duration of 72.7 ± 0.5 hours of data collection per person.

Due to the lower variability in the data obtained from the TT compared to the ST, this data set was used to calculate equivalent VO_2_ consumption based on HR, during occupation-related tasks. The average time spent in equivalent VO_2_ zones for the most strenuous session is represented in Table 3. There was a significant difference observed between the percentage of time spent in each VO_2_ zone (F = 16.293, p ≤ 0.001), where participants spent the majority of their time working at or below an oxygen demand of 20 ml/kg/min.

**Table 3.** Percentage of time spent in VO_2_max equivalent zones.

| **FO (n)** | **AVRO (n)** | **VO<sub>2</sub> equivalent zones (ml/kg/min)** | **Percentage of wearing time per zone during the most strenuous session (%)** | **Average duration spent in each zone during the most strenuous session (mean ± SD) hours:minutes:seconds** |
| --- | --- | --- | --- | --- |
| 2 | 16 | <5 | 4 | 00:28:12 ± 00:26:49 |
| 2 | 16 | 5-10 | 35 * | 02:59:58 ± 02:34:33 |
| 2 | 16 | 10-15 | 35 * | 02:25:01 ± 01:51:45 |
| 2 | 16 | 15-20 | 11 * | 00:29:42 ± 00:32:20 |
| 2 | 16 | 20-25 | 4 | 00:07:10 ± 00:05:31 |
|  | 16 | 25-30 | 4 | 00:06:30 ± 00:08:39 |
|  | 16 | 30-35 | 4 | 00:04:21 ± 00:03:52 |
|  | 16 | 35-40 | 2 | 00:01:36 ± 00:01:45 |

FO – firearms officer, AVRO – armed vehicle response officer.
\* Indicates a significant difference between time spent in between 5-20 ml/kg/min compared to above 20 (p < 0.05).

Of the 18 participants, all delivered firearms officers (FO) training, and 16 of these (15 male) also delivered armed vehicle response officer (AVRO) training. The 16 AVRO instructors reached oxygen demand levels beyond 30 ml/kg/min, only while training AVROs. Instructors who delivered FO training did not exceed an oxygen demand of 30 ml/kg/min during sessions, while instructors delivering AVRO training worked at higher intensities than 30 ml/kg/min but did not exceed 40 ml/kg/min.

## Discussion

This study identified the maximal oxygen demand (VO_2_max) obtained during two incremental tests (the 15m shuttle test and the Bruce treadmill test). The Bruce treadmill test (TT) VO_2_max was used to determine the level of cardiorespiratory exertion that firearms instructors undergo when delivering operational firearms training. The main findings of this study suggest that instructing firearms officers demands an oxygen consumption of up to 30 ml/kg/min, instructing armed vehicle response officers demands an oxygen consumption of up to 40 ml/kg/min, and that the 15m shuttle test (ST) is more physically demanding than the treadmill test(TT).

### Comparison of the shuttle test with the treadmill test

Optimal cardiovascular fitness is a key requirement for safeguarding health and promoting effective performance in law enforcement. The ST is a practical field-based test, typically administered with the assumption that specific stages of the test equate to estimated levels of VO_2_max [25,26]. In this study, the ST resulted in significantly higher peak oxygen demands compared to the TT. This falls in line with Morris et al [27] findings, where measured VO_2_max during the ST was higher than the estimations provided by Brewer [28] (Table 2).

This information indicates that the ST is a more strenuous test than the TT, where participants present a higher oxygen demand on the ST and reach exhaustion sooner. This is likely due to the higher physical requirements of constant changes in direction on the ST. The TT involves continuous linear running and is therefore primarily reliant on endurance qualities, while the ST requires regular changes of direction, adding a reliance on agility, eccentric forces to decelerate, and explosive concentric force to accelerate. The increased oxygen demand during the ST is therefore almost certainly due to the nature of musculature loading during deceleration and change of direction, with greater angles of change of direction requiring a greater muscle activation [26]. As a result, the change of direction increases the metabolic cost of task, resulting in higher cardiovascular requirements [29]. These increases in muscle activation and metabolic cost have been shown to be further impacted by an individual’s lower body strength and power [30]. Additionally, a 180-degree change of direction is further impacted by co-ordination and test familiarity, causing a high degree of variability in test outcome [25]. This is reflected in the results presented here, where there was no correlation between the VO_2_max obtained on the two tests, indicating that, although the ST consistently produced higher values, there was no defined ratio to this relationship. Therefore, ST might be best used to provide a VO_2_max predictive range rather than a definitive VO_2_max per stage reached [25,31]

Measuring VO_2_max on a treadmill presents a progressive test with less variability and less strain on the lower limbs, providing a more accurate estimate and reducing the risk of injury [17,18,32]. Therefore, if the intention is to purely obtain a measure of cardiovascular fitness, an ergometer test (eg. treadmill or bike) provides a more reliable direct measure of VO_2_max. If, however, the intention is to measure a more complex fitness ability which includes cardiovascular fitness, lower limb power, acceleration and agility, a shuttle test could be more appropriate.

This study concentrated purely on the cardiopulmonary demands of instructors delivering training to law enforcement officers, therefore, any mention of VO_2_ hereafter will be made in reference to values obtained through the TT, as this is the more reliable measure of cardiovascular fitness.

### Physical requirements of occupation-related tasks

FI spent most of their most strenuous day at or below 20 ml/kg/min. However, it should be noted that a significant amount of time (7:10 ± 5:30 minutes) was spent between 20-25 ml/kg/min. To be able to sustain this workload without the detrimental accumulation of fatigue, a minimum anaerobic threshold of 25 ml/kg/min could be recommended. Measuring an anaerobic threshold requires expensive equipment, while VO_2_max can be estimated using indirect tests. Given that the ratio of anaerobic threshold to VO_2_max in the general population is typically between 55 - 65% [33,34], a minimum VO_2_max between 38.5 – 45.5 ml/kg/min would therefore be required to sustain this level of effort without incurring excessive fatigue. This would put individuals in the best position to sustain the delivery of firearms training courses with minimal cardiovascular strain [34–37] It should also be considered that a “good” rating in cardiorespiratory fitness ranges from 36-56 ml/kg/min for 18-55-year-old females and 40-60 ml/kg/min for males (See ACSM, 2014 for a detailed table of normative values) [34]. Good cardiorespiratory function is crucial to safeguard general health and reduce the risk of coronary heart disease, myocardial infarction and overall mortality, where values below 28 ml/kg/min present a significantly greater risk of these incidents [35–37]. It is also worth noting that higher cardiorespiratory fitness is associated with improved cognitive function and motor processes through ageing [38], which also allows for greater control of the timing of shot with cardiac rhythm, improved shot success, decision-making, alertness and memory [7–12].

When delivering FO training, instructors did not exceed an oxygen demand of 30 ml/kg/min. FO training consisted of classroom-based lectures, firearms instruction on the firing range and assailant restraint and control training. AVRO training which consisted of FO training and the addition of open country assailant search and restraint as well as vehicle stop and restraint instruction was found to be more demanding. Instructors sustained a small but meaningful period (1:36 ± 1:45 minutes) between 35-40 ml/kg/min of equivalent VO_2_ during open country assailant search and restraint, which required FI to maintain instruction of officers whilst they are searching, chasing down and restraining an assailant in a simulated absconding drill, this approaches maximal intensities for some participants included in this study. Assuming FI should not be working at their maximal physical capacity while instructing others, it should be strongly recommended that these FI should hold a VO_2_max that exceeds this bracket.

Therefore, taking into consideration a required anaerobic threshold of 25 ml/kg/min, FI who only train FOs could be recommended to have a VO_2_max that exceeds 38.5 ml/kg/min. However, considering the significantly greater physical demands of AVRO training, instructors who train these officers could be recommended to have a VO_2_max that noticeably exceeds 40 ml/kg/min. It is important to note here that these measurements only relate to the physical demands placed on instructors while training other law enforcement officers, they do not relate to the fulfilment of real-life operational tasks, which are highly likely to exceed these demands.

### Cardiovascular exercise recommendations

Utilising a multi-modal approach of cardiovascular exercise prescription seems to suggest the greatest long-term yield for adaptation not only for the cardiovascular system but also for body composition [39–41]. This would include a comprehensive strategy to train physiological outputs that would improve centrally driven factors such as cardiac output and stroke volume [42–44]concurrently with stimuli that would cause peripheral adaptations such as mitochondrial biogenesis, upregulation of capillary density and A-VO_2_ difference, thus improving VO_2_max as well as reducing BMI and body fat percentage [39–44]. This involves the concurrent prescription of steady-state continuous effort exercise (SS), high-intensity interval training (HIIT) and sprint interval (SI) exercise. Evidence suggests that an 8-week 2-3x per week HIIT program will start to elicit cardiovascular central adaptations to improve VO_2_max but at the cost of reduced enjoyment and increased discomfort when compared to SS training [39,45]. This alone may put individuals off its completion, furthermore, peripheral and body composition adaptations were not seen until 12 weeks, aligned with the adaptations of SS training [39–45]. Alone HIIT or SI training did not display any greater physiological adaptations long-term when compared to SS, except for a minor increase in lean muscle mass for SI compared to SS[39]. Thus, the concurrent prescription of all three modalities across a 12+ week training program is postulated to simultaneously improve central, peripheral and body composition adaptations. This will also reduce training monotony, which is a major limiting factor for training participation [46]. Therefore, a 2-3x per week 12-week training program that mixes steady state, high intensity interval and sprint interval with a focus on long-term physiological and body composition adaptations, whilst keeping sessions enjoyable, should be emphasised to minimise long-term cardiorespiratory illness in the more senior FI.

### Limitations and further research

The sample size of the study was limited due to access to a small specialist population, where data was also lost due to participants being called on operation while being tested. This study focused on cardiorespiratory requirements only; it did not investigate other physical requirements needed to safeguard FI from injury during role-playing, resistance and restrain scenarios. Strength and power are key requirements for many armed operational tasks and should also be considered when assessing the physical burden of instructor and officer training. An enquiry into the prevalence of musculoskeletal injuries obtained during firearms drills could also provide an understanding of injury risk in this population, enabling the development of risk reduction strategies. It is worth noting that even though participants were instructed to refrain from caffeine and exercise, as well as arrive in a hydrated state this could not be controlled for and may have influenced their body composition assessment. Also, the use of different equipment to measure VO_2_max during testing may have influence on the final testing output. An estimation calculation could be used to predict VO_2_max, but there is no literature to validate its use in a 15m shuttle run test.

It should be underlined that these results apply to males but may not necessarily apply to females. The data set only included one female; even though this participant’s values matched the average values obtained by the wider male cohort, one person is not representative of the wider female population. Further research focused on female firearms instructors is required to ensure that recommended values are appropriate to either gender.

Finally, these results apply to instructors delivering training, but not to real-life operational tasks. A similar enquiry on cardiorespiratory demands of operational firearms tasks could provide valuable insight into the demands of specialist officers to inform fitness testing requirements in this cohort.

### Conclusions

A comparison of the oxygen demands of the treadmill test and shuttle test suggested that the shuttle test provides more variable results, due to greater demands of motor control, agility and explosive strength. Therefore, a treadmill test appears to provide a more accurate evaluation of cardiorespiratory fitness, while a shuttle test may provide a more global assessment of cardiorespiratory fitness, lower limb power and agility together. Using the outputs from the treadmill test to estimate cardio-pulmonary demands of occupational tasks, the results indicate that FI reached oxygen demands of 30 ml/kg/min when training AROs, however, this oxygen demand may reach 40 ml/kg/min when training AVROs. Therefore, FI requires high levels of cardiorespiratory fitness to withstand the physical demands of the task. This indicates that highly experienced FI should continue to maintain levels of fitness that exceed a VO_2_max of 40 ml/kg/min even after becoming non-operational, allowing the effective delivery of training and safeguarding their long-term health.

## Acknowledgements

We thank Justin Dixon for comments on the manuscript and Derek Brooks for organisation and logistics, making scheduling around operational shifts, work-related requirements, and facilities a harmonious process.

## Funding statement

This research received funding from a large British Law Enforcement Service, that remains unnamed due to a legally binding confidentially agreement. The funders had no role in study design, data collection and analysis, decision to publish, or preparation of the manuscript.

## Data availability statement

Data can be requested from and quote Project ID: 13985/004 or through the corresponding author upon reasonable request and in line with the legally binding confidentiality agreement between University College London and the counter-signed British Law Enforcement Service.

## Notes

### Competing Interest Statement

The authors have declared no competing interest.

### Funding Statement

Yes this research was funded under a confidentiality agreement with the Large British Police Authority.

### Author Declarations

The study was approved by UCL's Ethics Committee (Project ID number: 13985/004) in line with the declaration of Helsinki.

### Summary of Updates

Amended document according to some current reviewer comments and also to be in line with the confidentiality agreement in place with the Police Authority.

